# Humoral immune responses remain quantitatively impaired but improve qualitatively in anti-CD20 treated patients with multiple sclerosis after three or four COVID-19 vaccinations

**DOI:** 10.1101/2022.08.10.22278639

**Authors:** Carolin Otto, Tatjana Schwarz, Lara M. Jeworowski, Marie L. Schmidt, Felix Walper, Florence Pache, Patrick Schindler, Moritz Niederschweiberer, Andi Krumbholz, Ruben Rose, Christian Drosten, Klemens Ruprecht, Victor M Corman

## Abstract

**Background:** Humoral immune responses to COVID-19 vaccination are diminished in anti-CD20 treated patients with multiple sclerosis (pwMS). In healthy individuals, neutralizing antibodies against the SARS-CoV-2 Omicron variant are only detected after three COVID-19 vaccinations. It was hitherto unknown whether a third or fourth COVID-19 vaccination of anti-CD20 treated pwMS improves SARS-CoV-2 specific humoral immune responses, including neutralizing antibodies against Omicron.

**Methods:** Anti-CD20 treated pwMS vaccinated two (n=61), three (n=57) or four (n=15) times and healthy controls (n=10) vaccinated thrice were included in a prospective cohort study. Anti-SARS-CoV-2 spike S1 IgG and IgA levels, maturation of SARS-CoV-2 IgG avidity, neutralizing capacity against the SARS-CoV-2 Omicron BA.2 variant and SARS-CoV-2 specific T cell responses were analyzed.

**Results:** The proportion of anti-CD20 treated pwMS with detectable SARS-CoV-2 S1 IgG was similar after the second (31/61, 50.8%), third (31/57, 54.4%) and fourth (8/15, 53.3%) vaccination. In pwMS with detectable SARS-CoV-2 IgG, the proportion with high affinity antibodies increased from the second (6/31, 19.4%) to the third (17/31, 54.8%) and fourth (6/8, 75%) vaccination. While none (0/10) of the anti-CD20 treated pwMS vaccinated twice had Omicron specific neutralizing antibodies, 3/10 (30%) pwMS vaccinated thrice and 3/5 (60%) pwMS vaccinated four times generated Omicron specific neutralizing antibodies.

**Conclusion:** Although SARS-CoV-2 specific humoral immune responses remain quantitatively impaired, in those anti-CD20 treated pwMS who do develop SARS-CoV-2 antibodies, the functionality of SARS-CoV-2 antibodies, including neutralizing antibodies against Omicron, improves after three and four SARS-CoV-2 vaccinations, supporting current recommendations for one or two booster vaccination in anti-CD20 treated pwMS.

## Introduction

Anti-CD20 therapy in patients with multiple sclerosis (pwMS) is associated with worse courses of Coronavirus disease 2019 (COVID-19), caused by severe acute respiratory syndrome coronavirus-2 (SARS-CoV-2).^1^ While COVID-19 vaccinations strongly reduce the risk of severe SARS-CoV-2 infections^2,3^, anti-CD20 treated pwMS vaccinated twice have diminished SARS-CoV-2 antibody levels, avidity and neutralizing capacity, although SARS-CoV-2 specific T cell responses are preserved.^4,5^ Following the dissemination of the SARS-CoV-2 Omicron variant since December 2021^6^, it was shown that individuals vaccinated thrice, but not twice, against SARS-CoV-2 showed neutralizing antibodies against Omicron.^7,8^ However, whereas guidelines meanwhile recommend up to five SARS-CoV-2 vaccinations in anti-CD20 treated pwMS^9^, it was unknown whether anti-CD20 treated pwMS generate neutralizing antibodies against Omicron after three or four COVID-19 vaccinations. Here, we characterize humoral and cellular immune responses to SARS-CoV-2 in anti-CD20 treated pwMS vaccinated three or four times against SARS-CoV-2, including neutralizing antibodies against Omicron.

## Patients and Methods

### Patients

Patients were recruited between 5 March 2020 and 27 June 2022 and participated in a previously described^4^ ongoing prospective study of anti-CD20 treated pwMS at the MS outpatient clinic, Charité Campus Mitte, Charité – Universitätsmedizin Berlin (online supplemental eMaterials).

### Laboratory methods

Anti-SARS-CoV-2 spike subunit 1 (S1, Non-VOC) immunoglobulin (Ig)G and IgA, maturation of SARS-CoV-2 S1 IgG avidity, neutralizing capacity against the SARS-CoV-2 Omicron BA.2 variant and SARS-CoV-2 spike specific T cell responses were measured as previously described^4^ (online supplemental eMaterials).

## Results

Details of anti-CD20 treated pwMS vaccinated two (n=61), three (n=57) or four (n=15) times and healthy controls (HC, n=10) vaccinated thrice against COVID-19 are summarized in online supplemental eTable 1. All pwMS had been treated with at least one intravenous infusion of anti-CD20 therapy (ocrelizumab or rituximab) within a maximum of 18 months prior to the last COVID-19 vaccination. All COVID-19 vaccinations were administered after the start of anti-CD20 therapies.

### Anti-SARS-CoV-2 IgG levels

The proportion of anti-CD20 treated pwMS with detectable SARS-CoV-2 S1 IgG did not increase between the second (31/61, 50.8%, 95CI:38.6-62.9), third (31/57, 54.4%, 95CI: 41.6-66.6) and fourth (8/15, 53.3%, 95CI: 30.1-75.2) COVID-19 vaccination (Figure 1A). Accordingly, SARS-CoV-2 S1 IgG levels in anti-CD20 treated pwMS were similar after the second (median [IQR] 1.2 [0.1-5.7]), third (1.2 [0.2-5]) and fourth (1.2 [0.1-6.6]) vaccination. In contrast, 10/10 (100%, 95CI: 72.2-100) HC vaccinated thrice against SARS-CoV-2 developed SARS-CoV-2 S1 IgG. Median [IQR] SARS-CoV-2 S1 IgG levels of HC (9.0 [8.3-9.5]) were significantly higher than those of pwMS after the third (*p*=0.0001) and the fourth (*p*=0.003) vaccination (Figure 1A). Similar results were observed for anti-SARS-CoV-2 S1 IgA (online supplemental eFigure 1A).

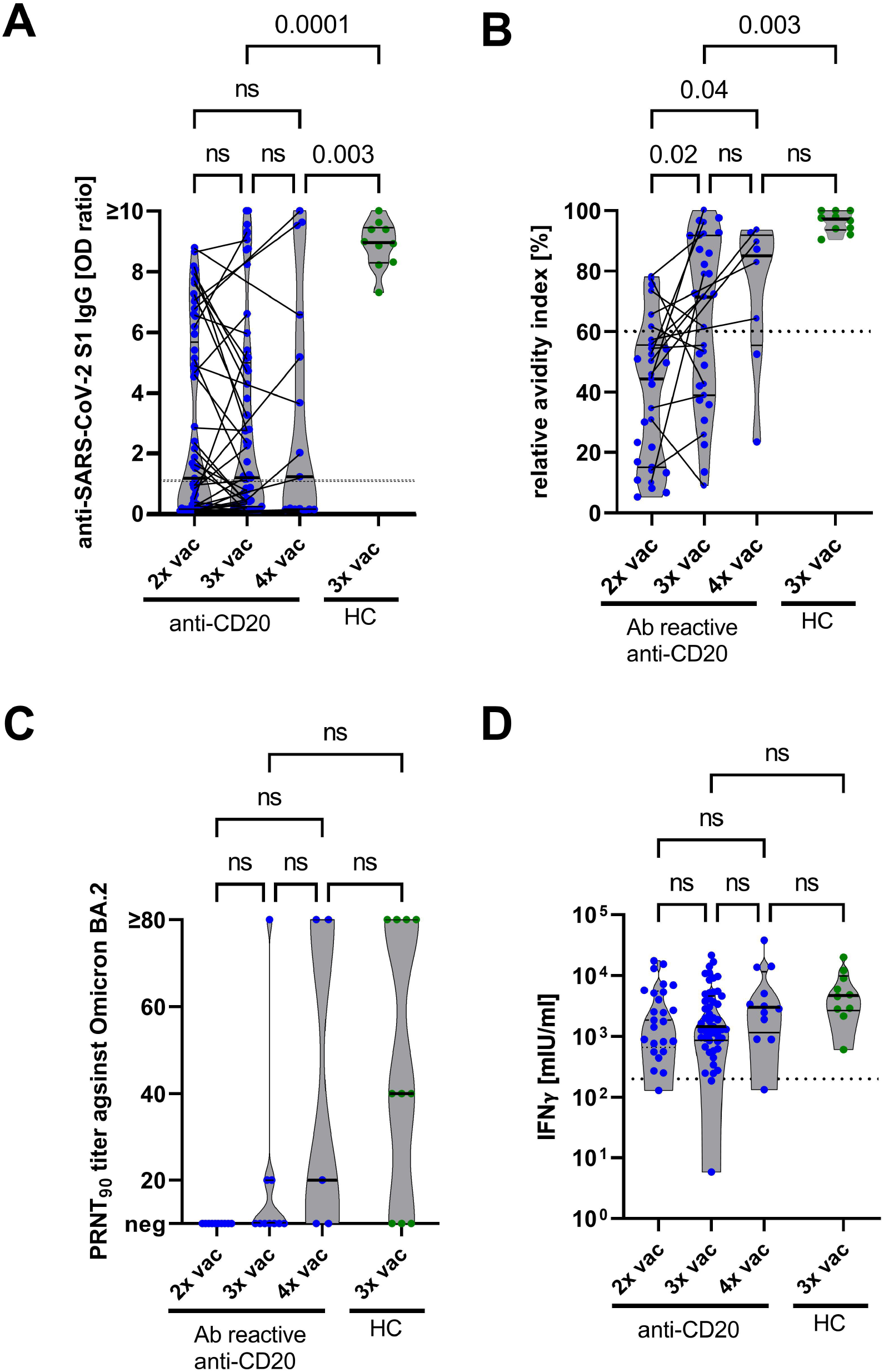
A. SARS-CoV-2 S1 IgG levels in two, three and four times vaccinated anti-CD20 treated pwMS and three times vaccinated HC were measured by ELISA. pwMS sampled more than once are connected by lines. The dotted horizontal line indicates the manufacturer’s threshold of an SARS-CoV-2 S1 IgG OD ratio of 1.1, levels above which were considered positive. B. Relative avidity indices in anti-S1 IgG reactive (Ab reactive) two, three and four times vaccinated anti-CD20 treated pwMS and three times vaccinated HC. pwMS sampled more than once are connected by lines. The dotted horizontal line indicates a relative avidity index of 60, levels equal to or above which were considered high. C. Serum neutralization titers against Omicron BA.2 were analyzed in two, three and four times vaccinated pwMS and three times vaccinated HC. Serum dilutions with a plaque reduction of 90% are referred to as PRNT90 titers. PRNT90 titers ≥20 were considered to indicate the presence of neutralizing antibodies. D. Whole blood of pwMS vaccinated two, three or four times and of HC vaccinated three times against SARS-CoV-2 was stimulated ex vivo with components of the S1 domain of the spike protein for 24 h, and interferon (IFN)-γ concentration in the supernatant was measured by ELISA. The dotted horizontal line indicates IFN-γ concentrations of 200 mIU/ml, levels above which indicate the presence of SARS-CoV-2 specific T cells. The pwMS with a very low T cell response following three vaccinations had an additional combined immunodeficiency syndrome. All *p*-values were calculated by the non-parametric Kruskal-Wallis test with Dunn’s multiple comparisons test. Abbreviations: HC=healthy controls, IFN-γ=interferon-γ, IgG=immunoglobulin G, IGRA=interferon-γ release assay, IU=international units, ns=not significant, OD=optical density, PRNT=plaque reduction neutralization test, S=SARS-CoV-2 spike protein S1 domain, vac=vaccination

Intraindividual courses of SARS-CoV-2 antibody responses could be studied in longitudinal blood samples collected after the second and third SARS-CoV-2 vaccinations from 35 pwMS (Figure 1A). 17/35 (48.6%, 95CI: 33-64.4) pwMS had detectable and 18/35 (51.4%, 95CI: 35.6-67) had undetectable SARS-CoV-2 S1 IgG after two vaccinations. Of the 18 pwMS without detectable SARS-CoV-2 S1 IgG after two SARS-CoV-2 vaccinations, only four (22.2%, 95CI: 9.0-45.2) showed detectable antibodies after the third vaccination, while 14 remained negative (77.8%, 95CI: 54.8-91). In five of 17 (29.4%, 95CI:13.3-53.1) pwMS with detectable SARS-CoV-2 S1 IgG after the second vaccination, SARS-CoV-2 S1 IgG was not detectable anymore after the third vaccination. In the remaining 12/17 (70.6, 95CI: 46.9-86.7) pwMS, SARS-CoV-2 S1 IgG stayed detectable also after the third vaccination.

SARS-CoV-2 S1 IgG levels after the fourth (*r*_*s*_=0.71, *p*=0.0036), but not after the third (*r*_*s*_=0.23, *p*=0.09, online supplemental eFigure 1B), vaccination increased with increasing time between the last anti-CD20 infusion and vaccination. The missing association of the interval between the last anti-CD20 infusion and the third vaccination might be related to a shorter median (IQR) interval between the last anti-CD20 infusion and the third vaccination (147 [122-183] days) as compared to the fourth vaccination (217 [155-247] days, *p*=0.05).

### Functionality of SARS-CoV-2 antibodies

Maturation of IgG avidity was analyzed in all anti-CD20 treated pwMS with detectable SARS-CoV-2 S1 IgG. After the second vaccination, only 6/31 pwMS (19.4%, 95CI: 9.2-36.3) exhibited high (≥60) SARS-CoV-2 S1 IgG avidity indices. However, after the third and fourth vaccinations, the proportion of pwMS with high SARS-CoV-2 IgG avidity indices increased to 17/31 (54.8%, 95CI: 37.8-70.8) and 6/8 (75%, 95CI: 40.9-95.6), respectively (Figure 1B). Accordingly, the median (IQR) avidity indices increased from the second (44.4 [IQR 15.1-55.7]) to the third (71.4 [39.0-91.8], *p*=0.02) and fourth (85.1 [55.5-91.9], *p*=0.04) vaccination. 10/10 (100%) HC had high median (IQR) avidity indices (97.7 (93.6-100).

The capacity of SARS-CoV-2 antibodies to neutralize Omicron (BA.2) was investigated using plaque reduction neutralization tests in pwMS with the highest SARS-CoV-2 S1 IgG levels after the second (n=10), third (n=10) and fourth (n=5) vaccinations (Figure 1C). While none (0/10, 0%, 95CI: 0-27.8) of the pwMS vaccinated twice had Omicron specific neutralizing antibodies, 3/10 (30%, 95CI: 10.8-60.3, *p*=0.21 vs. two vaccinations) pwMS vaccinated thrice and 3/5 pwMS (60%, 95CI: 39.7-89.2, *p*=0.02 vs. two vaccinations) vaccinated four times generated Omicron specific neutralizing antibodies. Likewise, 7/10 HC (70%, 95CI:39.7-89.2) vaccinated thrice had Omicron specific neutralizing antibodies, which tended to be higher (median [IQR] 40 [0-80]) than those of pwMS vaccinated three (0 [0-20]) or four (20 [0-80]) times.

### Cellular SARS-CoV-2 specific immune responses

24/25 (96%, 95CI: 80.5-99.8) pwMS vaccinated twice, 49/51 (96.1%, 95CI: 86.8-99.3) pwMS vaccinated thrice, 11/12 (91.7%, 95CI: 64.6-99.6) pwMS vaccinated four times and 10/10 HC (100%, 95CI: 72.3-100) had SARS-CoV-2 specific T cell responses, as detected by an SARS-CoV-2 spike specific interferon (IFN)-γ release assay (Figure 1D). IFN-γ concentrations were similar across all analyzed groups.

## Discussion

This study shows that the proportion of anti-CD20 treated pwMS with detectable SARS-CoV-2 S1 IgG and IgA antibodies after two, three and four vaccinations remains stable at approximately 50%. Only about 20% of anti-CD20 treated pwMS, who did not have SARS-CoV-2 S1 IgG after the second vaccination, developed SARS-CoV-2 S1 IgG after the third vaccination, which complies well with previous findings in anti-CD20 treated patients, who did not seroconvert after the second vaccination.^10–12^ Importantly, in some previously SARS-CoV-2 IgG positive pwMS, SARS-CoV-2 IgG became undetectable after the third vaccination, likely due to a waning SARS-CoV-2 antibody response over time^13^, which could not be overcome by the third vaccination. Altogether, these findings indicate a persistently impaired quantitative SARS-CoV-2 specific humoral immune response in anti-CD20 treated pwMS even after two booster vaccinations.

However, as an important novel finding, in those anti-CD20 treated pwMS, who did develop SARS-CoV-2 antibodies following SARS-CoV-2 vaccinations, the functionality (avidity and neutralizing capacity) of SARS-CoV-2 antibodies improved after a third or fourth vaccination. This suggests that in some anti-CD20 treated pwMS, the physiological mechanisms of antibody maturation and diversification are preserved, though at an attenuated level.

Maintained SARS-CoV-2 specific T cell responses after three and four vaccinations are consistent with findings after two vaccinations^4^, and appear relevant as vaccine-induced T memory cells cross-recognize Omicron variants.^14,15^

Limitations of this study are the rather small number of anti-CD20 treated pwMS vaccinated four times and the lack of absolute B cell counts.

In conclusion, humoral responses to SARS-CoV-2 remain quantitatively impaired in anti-CD20 treated pwMS after three or four vaccinations, while SARS-CoV-2 specific cellular immune responses are maintained. However, in those pwMS who develop SARS-CoV-2 antibodies, the functionality of SARS-CoV-2 antibodies improves after three or four vaccinations, including increased antibody avidity and generation of neutralizing antibodies against Omicron. These results support current recommendations to perform one or two booster vaccinations in anti-CD20 treated pwMS.

## Supporting information

online supplemental eFigure

## Data Availability

Data are available upon reasonable request.

## Data availability statement

Data are available upon reasonable request.

## Ethics statements

The study was approved by the ethical committee of Charité - Universitätsmedizin Berlin (EA2/152/21 and EA1/068/20).

## Acknowledgments

We thank Patricia Tscheak and Petra Mackeldanz for excellent assistance.

## Author Contributions

CO: study concept and design, acquisition, analysis, or interpretation of data, statistical analysis and drafting of the manuscript. TS: study concept and design, acquisition, analysis, or interpretation of data, statistical analysis and drafting of the manuscript. LMJ: acquisition, analysis, or interpretation of data, administrative, technical, or material support and critical revision of the manuscript for important intellectual content. MLS: acquisition, analysis, or interpretation of data, administrative, technical, or material support and critical revision of the manuscript for important intellectual content. FW: acquisition, analysis, or interpretation of data, administrative, technical, or material support and critical revision of the manuscript for important intellectual content. FP: acquisition, analysis, or interpretation of data, administrative, technical, or material support and critical revision of the manuscript for important intellectual content. PS: acquisition, analysis, or interpretation of data, administrative, technical, or material support and critical revision of the manuscript for important intellectual content. MN: acquisition, analysis, or interpretation of data, administrative, technical, or material support and critical revision of the manuscript for important intellectual content. AK: administrative, technical, or material support and critical revision of the manuscript for important intellectual content. RR: administrative, technical, or material support and critical revision of the manuscript for important intellectual content. CD: administrative, technical, or material support and critical revision of the manuscript for important intellectual content. KR: study concept and design, acquisition, analysis, or interpretation of data, statistical analysis, drafting of the manuscript, administrative, technical, or material support and study supervision. VMC study concept and design, acquisition, analysis, or interpretation of data, statistical analysis, drafting of the manuscript, administrative, technical, or material support and study supervision.

