## Supplementary material for "Humoral immune responses remain quantitatively impaired but improve qualitatively in anti-CD20 treated patients with multiple sclerosis after three or four COVID-19 vaccinations": online supplemental

### **eMaterials**

**Patients with multiple sclerosis and healthy controls**

Patients with multiple sclerosis (pwMS) included in this study were recruited between 5 March 2020 and 27 June 2022 and participate in a prospective observational single-center cohort study of anti-CD20 treated pwMS at the multiple sclerosis outpatient clinic, Charité Campus Mitte, Charité Universitätsmedizin Berlin, Germany, which was previously described in detail.^1^ Inclusion criteria for the present study were age ≥17 years, a diagnosis of relapsing-remitting MS (RRMS) or primary progressive MS (PPMS) according to the McDonald 2017 criteria ^2^, at least on intravenous infusion of anti-CD20 therapy (ocrelizumab or rituximab) within a maximum of 18 months prior to the last SARS-CoV-2 vaccination and at least two SARS-CoV-2 vaccinations administered after the start of anti-CD20 therapies. Any pwMS with breakthrough SARS-CoV-2 infections were excluded from the study. Anti-CD20 therapy infusion schemes were as previously described.^1^ Blood withdrawals for assessment of SARS-CoV-2 antibody levels, avidity and neutralizing capacity as well as for SARS-CoV-2 interferon-γ release assays were performed on the day of intravenous anti-CD20 therapy infusion before the anti-CD20 infusion was administered. Blood samples were processed as previously described.^1^ In case pwMS had blood withdrawn on more than one occasion after the last SARS-CoV-2 vaccination, the first blood sample after the last SARS-CoV-2 vaccination was utilized for measurements.

10 employees of the Department of Virology, Charité - Universitätsmedizin Berlin, vaccinated three times against SARS-CoV-2, were included as healthy controls (HC).

**Laboratory methods**

Vaccine-induced anti-SARS-CoV S1 IgG and IgA were detected by commercially available anti-SARS-CoV-2 S1 IgG and anti-SARS-CoV-2 S1 IgA ELISAs according to the manufacturer’s instructions (Euroimmun, Lübeck, Germany) and as previously described^1^.

Maturation of SARS-CoV-2 IgG avidity was measured by a modified anti-SARS-CoV-2 S1 IgG ELISA (anti-SARS-CoV-2 S1 IgG ELISA Kit, Euroimmun). Briefly, sera were diluted 1:101 with sample buffer and incubated on SARS-CoV-2 spike proteins precoated plates for 1h at 37°C. After a washing step, 200 µl urea or 200 µl PBS were added to the plates and incubated for 10 min. Following a washing step, 100 µl conjugate and 100 µl substrate were added according to the manufacturer’s instructions. After a stop solution step, the optical density (OD) at 450 nm was measured. Data was interpreted by the calculation of the relative avidity index utilizing the equation (OD of the urea treated sample/OD of the PBS treated sample) x100%. A relative avidity index equal or above 60% was considered high avidity.

To detect neutralizing activity in sera after vaccinations, a plaque reduction neutralization tests (PRNT) was performed as described before^3^ but with a Omicron BA.2 isolate. Serum dilutions causing plaque reductions of 90% (PRNT90) were recorded as titres.

SARS-CoV-2 spike specific T cell responses were monitored using a commercially available interferon-γ (IFN-γ) release assay (IGRA, Euroimmun) according to the manufacturer's instructions. An IFN-γ concentration >200 mIU/ml was considered reactive.

**Statistical analyses**

Categorical variables are presented as frequencies and percentages with 95% confidence intervals (CI) calculated using the Wilson procedure with a correction for continuity.^4^ Continuous variables are presented as median values and interquartile ranges (IQR). Group comparisons were performed by either Fisher’s exact test or Chi square tests, as appropriate, or nonparametric Mann-Whitney *U* test or Kruskal-Wallis test with Dunn’s multiple comparisons test as appropriate. Correlation of two datasets was calculated by Spearman’s method. *P* values of <0.05 were considered statistically significant. Because of the exploratory nature of this study no correction for multiple comparisons was performed. For statistical analysis, GraphPad PRISM (version 9.2.0) was used.

**eFigure 1A and B**

A. Anti-SARS-CoV-2 S1 IgA responses were measured in serum after the second, third and fourth COVID-19 vaccination in anti-CD20 treated pwMS and after the third COVID-19 vaccination in HC. pwMS sampled more than once are connected by lines. The dotted horizontal line indicates the manufacturer’s threshold of an SARS-CoV-2 S1 IgG OD ratio of 1.1, levels above which were considered positive. *P*-values were calculated by a non-parametric Mann Whitney U test or by a Kruskal-Wallis test with Dunn’s multiple comparisons test. B. Correlation of anti-SARS-CoV-2 S1 IgG OD ratio and the time interval from last anti-CD20 infusion to the third or fourth vaccination in anti-CD20 treated pwMS. Correlations were calculated by Spearman’s method.

Abbreviations: HC=healthy controls, IFN-γ=interferon-γ, IgA=immunoglobulin A, ns=not significant, OD=optical density, S1=SARS-CoV-2 spike protein S1 domain, vac=vaccination

**eTable 1. Demographic, clinical and treatment characteristics of patients with multiple sclerosis as well as demographic characteristics of healthy controls**

|  | Anti-CD20 treated patients with MS | | | Healthy controls (hospital employees) | *p* value^a^ |
| --- | --- | --- | --- | --- | --- |
|  | Vaccinated two times | Vaccinated three times | Vaccinated four times | Vaccinated three times |  |
| Number | 61 | 57 | 15 | 10 |  |
| Age, median (range), years | 40 (20-68) | 44 (20-67) | 45 (29-70) | 31.5 (26-44) | 0.035^b^ |
| female/male (% female) | 35/26 (57%) | 36/21 (63%) | 7/8 (47%) | 7/3 (70%) | 0.59^b^ |
| RRMS/PPMS (% PPMS) | 50/11 (18%) | 47/10 (18%) | 10/5 (33%) | - | 0.37 |
| EDSS, median (range) | 2 (0-7.0) | 2 (0-7.0) | 3 (0-6.0) | - | 0.59 |
| Any previous immunotherapy before anti-CD20 therapy, n/total  n (%) | 33/61 (54%) | 34/57 (60%) | 6/15 (40%) | - | 0.78 |
| At least two previous immunotherapies before anti-CD20 therapy, n/total n (%) | 21/33 (63%) | 18/34 (53%) | 5/6 (83%) | - | 0.75 |
| Cumulative dose ocrelizumab per patient at latest blood withdrawal, median (range), mg | 1800 (600-4800) | 2400 (600-7200) | 3000 (0-7800)^c^ | - | - |
| Treatment with rituximab before ocrelizumab, n/total n (%) | 7/61 (12%) | 10/57 (18%) | 3/15 (20%) | - | - |
| Cumulative dose rituximab per patient, median (range), mg | 3000 (2000-5000) | 4000 (2000-9000) | 4000 (3000-9000)^c^ | - | - |
| Time from MS diagnosis to first sample collection within this study, median (range), months | 38 (1-334) | 41 (1-345) | 61 (3-272) | - | 0.75 |
| Applied vaccines | BNT162b2: n = 53  mRNA-1273: n = 4  ChADOx1 nCoV-19: n = 1  ChADOx1 nCoV-19/ BNT162b2: n = 3 | BNT162b2: n = 38  mRNA-1273: n = 2  BNT162b2/mRNA-1273: n = 13  ChADOx1 nCoV-19/ BNT162b2: n = 4 | BNT162b2: n = 6  BNT162b2/ mRNA-1273: n = 7  ChADOx1 nCoV-19/mRNA-1273: n = 2 | BNT162b2: n = 2  BNT162b2/ mRNA-1273: n = 1  ChADOx1 nCoV-19/BNT162b2 : n = 3  ChADOx1 nCoV-19/BNT162b2/mRNA-1273 : n = 4 |  |
| interval from last vaccination to blood withdrawal, median (IQR), days | 41 (33-140) | 52 (35-162) | 33 (26-133) | 30.5 (28.8-37) | 0.0006 |

^a^Statistical significance of differences in the age of the four groups, as well as of disease duration and EDSS in the three groups of vaccinated patients with MS (pwMS) was assessed by Kruskal-Wallis test; statistical significance of differences in the distribution of women/men among the four groups was assessed by Chi-square test, statistical significance of differences in the distribution of MS type, any previous immunotherapy and at least two previous immunotherapies was assessed by Fisher’s exact test.

^b^In group comparison with Fisher’s exact test, the distribution of women/men between the three MS groups (*p*=0.5) and between all four groups (p=0.59) did not differ. Comparison of age, assessed by Kruskal-Wallis test, revealed differences for all four groups (p=0.035), but not between the three MS groups (p=0.5). In pairwise group comparison with Mann-Whitney-U test, healthy controls were younger than all 85 pwMS combined (*p*=0.008).

^c^One patient had previously been treated with a cumulative lifetime dose of 9000 mg rituximab and anti-CD20 therapy was re-initiated with ocrelizumab within 18 months after the last rituximab treatment, sample collection for assessment was performed on the day of first ocrelizumab treatment, before drug administration.

EDSS = Expanded Disability Status Scale, MS = multiple sclerosis, n = number, RRMS = relapsing remitting multiple sclerosis, PPMS = primary progressive multiple sclerosis, BNT162b2 = Pfizer Inc. and BioNtech vaccine, mRNA-1273 = Moderna Inc. vaccine, ChADOx1 nCoV-19 = Oxford-AstraZeneca vaccine
