## Supplementary figures and images for "Humoral immune responses remain quantitatively impaired but improve qualitatively in anti-CD20 treated patients with multiple sclerosis after three or four COVID-19 vaccinations"

### online supplemental eFigure

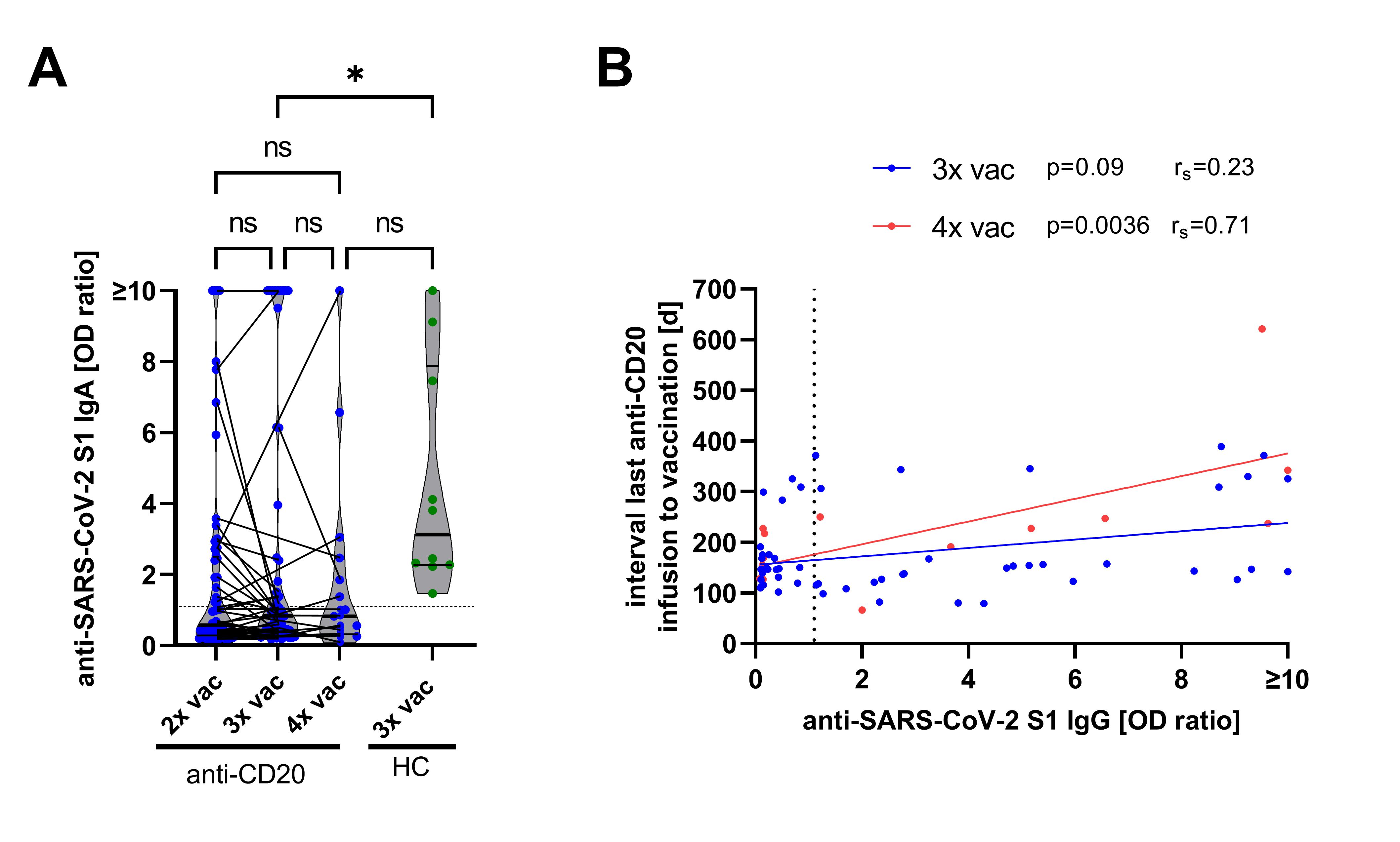
